# The Efficacy and Safety of Leflunomide in the Treatment of Giant Cell Arteritis: A Protocol for a Systematic Review

**DOI:** 10.1101/2025.07.06.25330932

**Authors:** Linda M. Zhu, Arielle Mendel, Carolyn Ross, Jean-Paul Makhzoum

**Affiliations:** Hopital Sacre-Coeur, University of Montreal, 5400 Bd Gouin O, Montreal, Qc, H4J1C5, Canada; Vasculitis and Lupus Clinic, McGill University Health Center, McGill University, Canadian Vasculitis Research Network, 1001 Bd Decarie, Montréal, QC H4A 3J1, Canada; Vasculitis Clinic, Hopital Sacre-Coeur, University of Montreal, Canadian Vasculitis Research Network, 5400 boul Gouin O, Montreal, Qc, H4J1C5, Canada; Vasculitis Clinic, Hopital Sacre-Coeur, University of Montreal, Canadian Vasculitis Research Network, 5400 Bd Gouin O, Montreal, Qc, H4J1C5, Canada

**Author notes:** **Correspondence**: Linda M. Zhu Vasculitis Clinic, Hopital Sacre-Coeur, University of Montreal, 5400 Bd Gouin O, Montreal, Qc, H4J1C5, Canada.

**Keywords:** Giant cell arteritis, Temporal arteritis, Leflunomide

## Abstract

**Background:** Giant cell arteritis (GCA) is a type of large vessel vasculitis that causes inflammation, scarring and stenosis in large vessels. Clinical symptoms include headaches, visual changes and myalgias. The standard treatment for GCA consists of glucocorticoid therapy, however long-term use of glucocorticoid therapy poses important health risks. For this reason, there has been a growing interest in exploring other glucocorticoid-sparing therapies, such as leflunomide. The objective of this systematic review is to assess the efficacy and safety of leflunomide in the treatment of GCA.

**Methods:** We will include studies with adult patients who received leflunomide in the treatment of GCA. We will include randomized controlled trials, cohort studies, case-control studies, and case series. The intervention of interest is the use of leflunomide, which may be initiated at any time over the course of the disease. The comparator is glucocorticoid therapy alone. The primary efficacy outcome is the incidence (proportion) of patients who attain GC-free remission, as defined by the following: 1) absence of signs or symptoms of GCA, and/or 2) normalization of inflammatory markers, and/or 3) radiologic response, and 4) complete discontinuation of GC. Outcomes will be assessed after 1 year of follow-up. Quality appraisal will be performed using the respective risk of bias tools. We will perform a meta-analysis using a random-effects model if the included studies are sufficiently homogenous. For quantitative synthesis, we will employ Cochran-Mantel-Haenszel method with odds ratios as a measure of effect and 95% confidence intervals for our primary endpoint.

**Discussion:** The challenge in the treatment of GCA is attaining and maintaining disease remission with the least amount of glucocorticoid therapy possible. The only approved glucocorticoid-sparing agent in the treatment of large vessel vasculitis is tocilizumab. Leflunomide is a molecule with similar downstream effects on dendritic cells and cytokines as tocilizumab. Should leflunomide demonstrate similar benefits, it would be a cost-effective, safe, and user-friendly (oral route) option.

**Systematic review registration:** Our systematic review protocol was registered with the International Prospective Register of Systematic Reviews (PROSPERO, registration number CRD42023490373).

## Background

Giant cell arteritis (GCA) is a type of large vessel vasculitis, where inflammatory processes cause scarring and stenosis in large vessels. It is a disease that is more common in patients aged 50 years and older (1) and is more likely to affect temporal arteries.(2) Clinical symptoms at time of presentation include headaches, scalp tenderness, jaw claudication, visual changes (including amaurosis fugax), and polymyalgia rheumatica.

Treatment for GCA consists of glucocorticoid (GC) therapy with early initiation of high dose therapy for induction, following with slow tapering over a year according to European League Against Rheumatism (EULAR).(3, 4) Despite this, there is a high rate of relapse during GC tapering or cessation, in approximately 30-80% of patients.(5-7) In a recent systematic review, the prevalence and incidence of major relapse was estimated to be 3.3% and 14.5/100 patient-years respectively.(8) Furthermore, long-term use of GC therapy poses important risks for diabetes, osteoporosis, infections and cardiovascular disease.(9) For this reason, in recent years, there has been a growing interest in exploring other non-GC therapies.

To date, tocilizumab, a monoclonal anti-interleukin-6 receptor antibody, has shown efficacy in GCA.(10, 11) However, tocilizumab is not without important risks and side effects, such as severe infections and leucopenia.(10, 11) Leflunomide is a dihydroorotate dehydrogenase inhibitor, with downstream effects on dendritic cells with inhibitory effects on cytokines such as IL-1, IL-6 and IL-17.(15) Thus, growing interest in leflunomide in GCA is explained by its low cost, favorable safety profile and ease of use (orally).

The objective of this systematic review is to assess the efficacy and safety of leflunomide as a GC sparing agent in the treatment of new-onset, refractory or relapsing GCA.

## Methods

### Research Questions

The proposed systematic review will aim at answering the following questions:

1. In adult patients with new-onset, refractory or relapsing GCA, does the addition of leflunomide to GC therapy, compared to GC therapy alone, improve remission rate, reduce relapse and GC usage?
2. In adult patients with new-onset, refractory or relapsing GCA, does the addition of leflunomide to GC therapy, compared to GC therapy alone, reduce adverse events?

### Outcomes

The primary efficacy outcome is the incidence (proportion) of patients who attain sustained GC-free remission, as defined by the following criteria:

- Absence of signs or symptoms of GCA (i.e. resolution of headache, jaw claudication, hip or girdle pain/stiffness, vision impairment, scalp tenderness, constitutional symptoms), and/or
- Normalization of inflammatory markers (i.e. C-reactive protein, erythrocyte sedimentation rate), and/or
- Radiologic response on ultrasound or ^18^F-FDG PET, and
- Complete discontinuation of GC.

The secondary efficacy outcome is the mean daily GC dose in patients taking leflunomide, compared to patients on prednisone alone.

The primary safety outcome is the incidence (proportion) of adverse events during treatment. We will extract outcomes in the measures of association as they are reported in the studies.

Outcomes will be assessed after 1 year of follow-up, or at 6 months for studies of shorter duration.

### Eligibility Criteria

We will include studies with adult patients (18 years or older) who received leflunomide in the treatment of new-onset, refractory or relapsing GCA. The diagnosis of GCA is made based on the American College of Rheumatology classification criteria (version 1990 or version 2022).(17, 18) New-onset GCA is defined a new diagnosis of GCA in a participant without a history of GCA. Refractory GCA is GCA that is persistent despite adequate GC therapy or flares up with GC dose reduction. Relapsing GCA is defined as active GCA in a participant who was previously in remission (i.e. asymptomatic GCA). Studies on other vasculitides will be excluded. Leflunomide doses of 10 mg daily with possibility of escalating to up to 20 mg daily will be included in the study. Leflunomide may be initiated at any time over the course of the disease, whether at first diagnosis of GCA, in relapsing or refractory disease.

We will exclude studies where the comparator group involves any other form of treatment that is not GC therapy alone.

We will include randomized controlled trials (RCTs), cohort studies, case-control studies, and case series. We will exclude cross-sectional studies, literature reviews, and case reports. At least 6 months of study duration will be required for inclusion. There will be no restrictions on the language of publication or on the setting of the study (inpatient, outpatient, academic or community hospital).

### Search Sources

The following databases will be searched: MEDLINE, Cochrane Central Register of Controlled Trials in the Cochrane Library (CENTRAL) and EMBASE. Study registries namely clinicaltrials.gov will also be searched. Grey literature will be searched, including conference papers, abstracts and presentations, namely from the American College of Rheumatology Annual Meeting and the European League against Rheumatism Annual Meeting.

A list of experts will be generated based on references from included studies. We will perform a citation index search of these experts on “Web of Science” and “researchgate.net” to ensure all potentially relevant studies are included.

We will also search study retraction or errata statements for all included studies to document inconsistencies following publication of study results.

### Search Strategy

Search strategies will be developed using medical subject headings (MeSH) and text words pertaining to GCA and leflunomide. A publication date filter will be applied from August 1^st^, 1990 to onwards, as the classification criteria were first published in August 1990. There will be no language restriction or any other filters.

The search strategy was developed by JPM and a Health Sciences Librarian and was reviewed by all co-authors. The transcript for MEDLINE can be found in Table 1. All other search strategies can be found in Supplementary Material. The search date will be provided at the review stage.

### Study Records Management, Selection, and Collection

All literature search results will be exported into Covidence (covidence.org), an online systematic review data management software. Duplicates from the searches will automatically be removed before the initial screening by Covidence. For studies with multiple reports, one record will be considered as the main source and this choice will be justified. Prior to beginning the official study selection process, two reviewers (LZ, JPM) will independently screen 10 records as a calibration exercise. A third reviewer will be available to resolve discrepancies within the Covidence software as needed.

Full text will be obtained for studies meeting inclusion criteria. Full-text screening will be performed by the same two reviewers. Reason for exclusion in full text will be documented and any disagreement will be resolved through discussion and/or by the third reviewer.

The two reviewers will then collect data independently using Covidence. Variables of interest include: study identifying information (study ID, record ID), reason for exclusion, characteristics of the study (author, year, country, design, duration, funding, conflict of interest), characteristics of participants (age, sex, ethnicity, baseline characteristics relating to GCA), details of the intervention (time to leflunomide initiation, indication for leflunomide, dose of leflunomide, dose of GC at time of leflunomide initiation), details of the comparator (dose of GC and/or other immunosuppressive therapy), outcomes (definitions, measures of association, timing), and results (number of participants, exclusions, losses at follow-up, summary results, subgroup results). Key conclusions, comments and references will also be collected.

We will document missing data as they are recorded. Study authors will be contacted for missing data or clarifications via email. A maximum of 3 attempts will be made, with follow-up emails every 2 weeks, in the case of no response.

### Quality Appraisal

Quality appraisal will be performed using a risk of bias tool according to the type of study. We will use version 2 of the Cochrane risk-of-bias tool for randomized trials (RoB 2) for randomized studies.(19) The “Risk Of Bias In Non-randomised Studies - of Interventions” (ROBINS-I) tool will be used to appraise risk of bias for non-randomised intervention studies,(20) while the “Risk Of Bias In Non-randomised Studies - of Exposure” (ROBINS-E) tool will be used to assess risk of bias for observational studies.(21)

The methodological quality assessment will be conducted by two independent reviewers (LZ, JPM). Discrepancies would be resolved by discussion and if needed, by a third reviewer. All findings will be summarized in a risk-of-bias table and figure using the RevMan software.

### Data Synthesis

We will perform a meta-analysis using a random-effects model if the included studies are sufficiently homogenous in terms of participants (indications for initiation of leflunomide), interventions (doses and duration of leflunomide), comparator and study design.

Our data will be mostly binary: proportions of GC-free remission, low-GC sustained remission and adverse events. For quantitative synthesis, we will employ Cochran-Mantel-Haenszel method with odds ratios (OR) as a measure of effect and 95% confidence intervals.

For our secondary endpoint of mean reduction in prednisone dose, we will use the mean difference as our measure of association. We will use the inverse variance method for quantitative synthesis. If there is ≥10% missing data for any outcome, we will perform a sensitivity analysis to assess its impact.

We will test statistical heterogeneity using Chi^2^ test (significance level of 0.1) and I^2^ statistic. If there is a high level of statistical heterogeneity (*p*<0.1 or I^2^ > 50%), we will analyse clinical heterogeneity by examining the variability in participants, interventions, and outcomes in the included trials. We will also compare study designs and settings to assess methodological heterogeneity.

Sensitivity analyses will be performed by excluding studies deemed to have a high risk of bias according to the Cochrane RoB 2.0, ROBINS-I and ROBINS-E tools.

We will also perform subgroup analyses based on disease subtype (new-onset vs. relapsing).

If a quantitative analysis is deemed inappropriate, we will provide a systematic narrative synthesis. Information will be presented and summarized in text and tables, with emphasis on outcomes of interest and subgroups. Studies at high risk of bias will be summarized in a separate table.

### Meta-Bias Assessment

If we include more than 10 studies in the final analysis, a funnel plot will be provided. We will use Egger’s test for binary outcomes to assess publication bias.(22)

Outcome reporting bias will be assessed by comparing the reported outcomes in the study with planned outcome measures in the same study’s protocol. Outcome reporting bias will be considered low if the following two conditions are met: 1) study protocol was published before availability of study results and 2) every ‘a priori’ outcome in the protocol is reported in the study or if it is not reported, then a justification is provided in the body of the study. If the study protocol is not available, we will use the Outcome Reporting Bias in Trials (ORBIT) tool to evaluate outcome reporting bias.(23)

A sensitivity analysis will be conducted if one or more studies present a high risk of outcome reporting bias.

### Confidence in the Evidence

Two reviewers will independently appraise the quality of evidence for outcomes in accordance to the Grading of Recommendations, Assessment, Development and Evaluation working group methodology (GRADE).(24) The GRADE assessment will be presented in a table with an overall GRADE assigned to all the outcomes in general.

## Discussion

One of the challenges in the treatment of GCA is attaining and maintaining disease remission with the least cumulative GC doses. The only approved GC-sparing agent is tocilizumab.

However, GCA relapses still occur with tocilizumab. Moreover, weekly doses of tocilizumab produce an estimated adjusted incremental cost-utility ratio to be $187,389 per quality-adjusted life-year.(25) If leflunomide shows efficacy in the treatment of GCA, it would provide a cost-effective therapeutic alternative.

The strength of this review lies in its methodology. We focus specifically on leflunomide and exclude all studies with comparators that are not GC alone. We chose clinically relevant endpoints. Our definition of the primary outcome includes clinical, laboratory and radiological findings which are often reported in a heterogeneous manner in the literature but remains robust in terms of the objective endpoint with complete discontinuation of GC. We also include grey literature in our search strategy. We anticipate some difficulties with our secondary efficacy outcome. A reported cumulative GC dose would have been a more precise tool in terms of assessing the GC sparing ability of leflunomide, but due to reporting convention in studies, we opted for the mean daily GC dose instead.

We aim to publish the results of this study in a peer-reviewed journal. Any amendments to the protocol will be reflected on PROSPERO and in the final manuscript.

## Data Availability

There are no new data associated with this article.

## List of Abbreviations

ACR: American College of Rheumatology
CENTRAL: Cochrane Central Register of Controlled Trials.
EULAR: European League Against Rheumatism
GC: Glucocorticoid
GCA: Giant cell arteritis
GRADE: Grading of Recommendations, Assessment, Development, and Evaluations
MeSH: Medical Subject Headings
OR: Odds ratio
ORBIT: Outcome Reporting Bias in Trials
PROSPERO: International Prospective Register of Systematic Reviews
RCT: Randomised controlled trial
RoB: Risk of bias
ROBINS-I: Risk of Bias in Non-randomised Studies – of Interventions
ROBINS-E: Risk of Bias in Non-randomised Studies – of Exposure

## Declarations

### Ethics approval and consent to participate

This study does not contain any research conducted on human participants or animals that require institutional ethics review approval.

### Consent for publication

Not applicable.

### Availability of data and materials

Not applicable.

### Authors’ contributions

LZ and JPM drafted the protocol and manuscript. All authors contributed to the development of the selection criteria, the risk of bias assessment strategy and data extraction criteria. LZ and JPM developed the search strategy. CR, JPM and AM provided expertise on giant cell arteritis. All authors read, provided feedback, and approved the final manuscript. JPM is the guarantor.

## Acknowledgements

Mrs. Guylaine Marcotte (research coordinator) and Dr Alexandra Mereniuk (collaborator).

---

**Figure 1.**
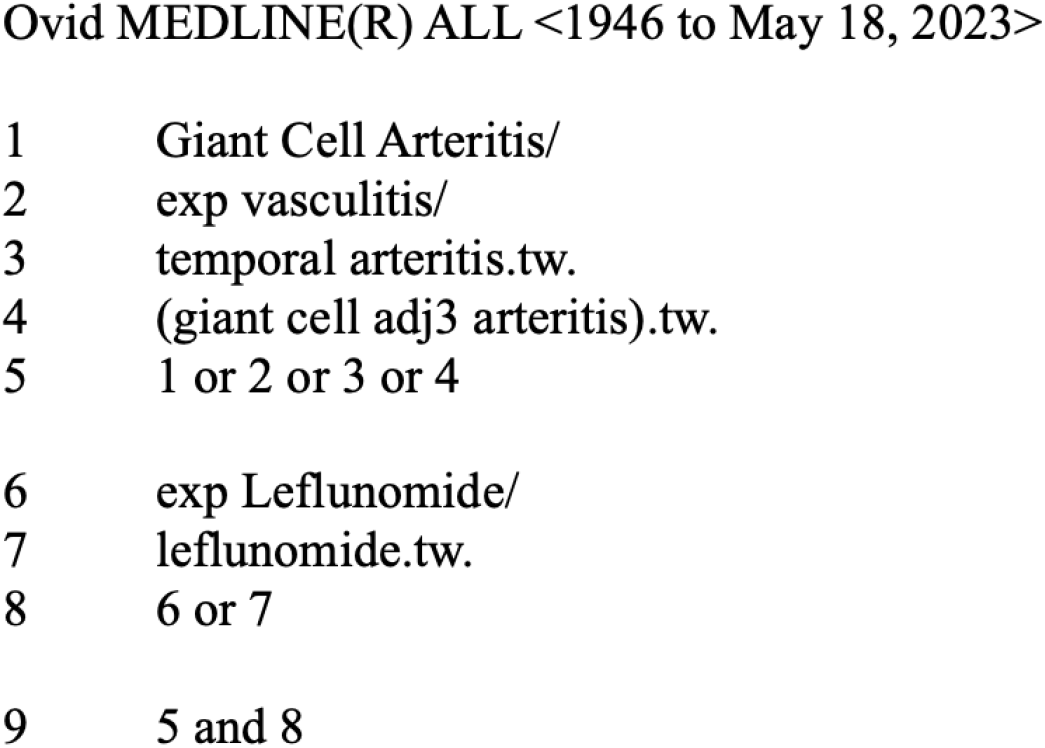
MEDLINE (OVID) search strategy

